# Sex differences in immune responses to SARS-CoV-2 that underlie disease outcomes

**DOI:** 10.1101/2020.06.06.20123414

**Authors:** Takehiro Takahashi, Patrick Wong, Mallory K. Ellingson, Carolina Lucas, Jon Klein, Benjamin Israelow, Julio Silva, Ji Eun Oh, Tianyang Mao, Maria Tokuyama, Peiwen Lu, Arvind Venkataraman, Annsea Park, Feimei Liu, Amit Meir, Jonathan Sun, Eric Y. Wang, Anne L. Wyllie, Chantal B.F. Vogels, Rebecca Earnest, Sarah Lapidus, Isabel M. Ott, Adam J. Moore, Arnau Casanovas-Massana, Charles Dela Cruz, John B. Fournier, Camila D. Odio, Shelli Farhadian, Nathan D. Grubaugh, Wade L. Schulz, Albert I. Ko, Aaron M. Ring, Saad B. Omer, Akiko Iwasaki, the Yale IMPACT research team.

## Abstract

A growing body of evidence indicates sex differences in the clinical outcomes of coronavirus disease 2019 (COVID-19)^1-4^. However, whether immune responses against SARS-CoV-2 differ between sexes, and whether such differences explain male susceptibility to COVID-19, is currently unknown. In this study, we examined sex differences in viral loads, SARS-CoV-2-specific antibody titers, plasma cytokines, as well as blood cell phenotyping in COVID-19 patients. By focusing our analysis on patients with mild to moderate disease who had not received immunomodulatory medications, our results revealed that male patients had higher plasma levels of innate immune cytokines and chemokines including IL-8, IL-18, and CCL5, along with more robust induction of non-classical monocytes. In contrast, female patients mounted significantly more robust T cell activation than male patients during SARS-CoV-2 infection, which was sustained in old age. Importantly, we found that a poor T cell response negatively correlated with patients’ age and was predictive of worse disease outcome in male patients, but not in female patients. Conversely, higher innate immune cytokines in female patients associated with worse disease progression, but not in male patients. These findings reveal a possible explanation underlying observed sex biases in COVID-19, and provide important basis for the development of sex-based approach to the treatment and care of men and women with COVID-19.

## Main

Severe acute respiratory syndrome coronavirus-2 (SARS-CoV-2) is the novel coronavirus first detected in Wuhan, China, in November 2019, that causes coronavirus disease 2019 (COVID-19)^5^. On March 11th 2020, the World Health Organization declared COVID-19 a pandemic, and as of June 4th, cases are over 6.5 million globally, with more than 380,000 deaths attributed to the virus^6^. A growing body of evidence reveals that male sex is a risk factor for a more severe disease, including death. Indeed, globally, ∼60% of deaths from COVID-19 are reported in men^7,8^, and a recent cohort study of 17 million adults in England reported a strong correlation between male sex and risk of death from COVID-19 (hazard ratio 1.99, 95% confidence interval 1.88-2.10)^8^.

Past studies have demonstrated that sex has a significant impact on the outcome of infections and has been associated with underlying differences in immune response to infection^9,10^. For example, epidemiological data indicate that prevalence of hepatitis A and tuberculosis are significantly higher in men compared with women^11^. Also, viral loads are consistently higher in male patients with hepatitis C virus (HCV) and human immunodeficiency virus (HIV)^12,13^. Conversely, women mount a more robust immune response to vaccines^14^. These findings collectively suggest a more robust ability among women to control infectious agents. However, the mechanism by which SARS-CoV-2 causes more severe disease in male patients than in female patients remains unknown.

To elucidate the differential immune response against SARS-CoV-2 infection in men and women, we performed detailed analysis on the sex differences in immune phenotype via the assessment of viral loads, SARS-CoV-2 specific antibody levels, plasma cytokines/chemokines, and blood cell phenotypes.

## Overview of the study design

Patients who were admitted to the Yale-New Haven Hospital between March 18th and May 9th, 2020 and were positive for SARS-CoV-2 by RT-PCR from nasopharyngeal and/or oropharyngeal swabs were enrolled through the IMPACT biorepository study. Among total 198 patients enrolled in IMPACT study this period, we obtained freshly-isolated peripheral blood mononuclear cells (PBMCs), plasma, nasopharyngeal swabs or saliva samples from in total 93 patients for the present study. The detailed demographics and clinical characteristics of study participants are shown in Extended Data Table 1.

Nasopharyngeal swabs and saliva samples for virus RNA assessment along with blood samples were collected on the day of enrollment. Plasma and PBMCs were isolated from whole blood and plasma was used for titer measurements of SARS-CoV-2 spike S1 protein specific IgG and IgM antibodies (anti-S1 IgG and IgM) and cytokine/chemokine measurements. Freshly isolated PBMCs were stained and analyzed by flow cytometry analyses. A subset (n = 54) of study participants donated blood, nasopharyngeal swabs, and saliva longitudinally (information found in Extended Data Table 1). To compare the immune phenotype between sexes, two sets of data analyses were performed in parallel, baseline and longitudinal. As a control group, COVID-19 uninfected health care workers (HCWs) from Yale-New Haven Hospital were enrolled. Demographics and background information for the HCW group are found in Extended Data Table 1 and the demographics of HCWs for cytokine assays and flow cytometry assays for the primary analyses (main figures) are found in Extended Data Table 2.

### Baseline Analysis

The baseline analysis was performed on samples from the first time point of patients who met the following criteria: not in intensive care unit (ICU), had not received tocilizumab (Toci), and had not received high dose corticosteroids (CS; prednisone equivalent > 40 mg) prior to the first sample collection date. This patient group, Cohort A, consisted of 39 patients (17 men and 22 women). Patients on hydroxychloroquine and remdesivir were not excluded, and 29 and 3 patients were on these drugs at the first time point sample collection, respectively. The demographics and background clinical information of each patient are found in Extended Data Table 2 and 3, respectively. The main figures represent analyses of baseline values obtained from patients in Cohort A.

### Longitudinal Analysis

As a parallel secondary analysis, we performed longitudinal analysis on a total patient cohort (Cohort B). Cohort B included all patient samples from Cohort A (including multiple time point samples from the Cohort A patients) as well as additional 54 patients who did not meet the inclusion criteria for Cohort A. Pregnant patients, patients on active/recent chemotherapy for cancers, patients with metastatic cancers, patients who have had renal transplant, and patients with autoimmune/inflammatory diseases were not included. This analysis included multiple time point samples from 93 participants in total. Data from Cohort B were analyzed for sex differences in immune responses among patients using longitudinal multivariable analyses to control for age, treatment (Toci and CS), days from symptom onset and ICU status. Both adjusted least square means difference over time in immune response between male and female COVID-19 patients (Extended Data Tables 4) and adjusted least square means difference over time in immune response between male and female COVID-19 patients and male and female healthcare workers (Extended Data Tables 5) were performed.

For both baseline analysis and longitudinal analysis, types of samples available from each patient were variable, and information on the sample size (n) for each assay is shown in the respective figure legends or tables. For Cohort A, sample types obtained from each patient at the first time point can be found in Extended Data Table 3.

## Male patients have higher levels of key innate immune cytokines

We first compared the virus RNA concentrations of male and female patients. For Cohort A, the median concentrations of virus RNA assayed with nasopharyngeal swabs and saliva were both higher in males, but there was no significant difference by sex. Analysis of the viral load from Cohort B did not reveal a difference, either, although median virus RNA concentrations both in nasopharyngeal swab and saliva samples were again higher in males (Extended Data Table 4).

We found induction of anti-SARS-CoV-2 S1 protein specific IgG and IgM (anti-S1-IgG and –IgM) antibodies in the plasma of many patients, and IgG levels tended to be higher in female patients, though not significantly different from their male counterpart (Fig. 1b). In the analysis of Cohort B, the difference between sexes were not obvious (Extended Data Table 4 and 5). Thus, at baseline and during disease course, there are no clear differences in the amount of IgG or IgM generated against the S protein between male and female.

**Fig. 1.**
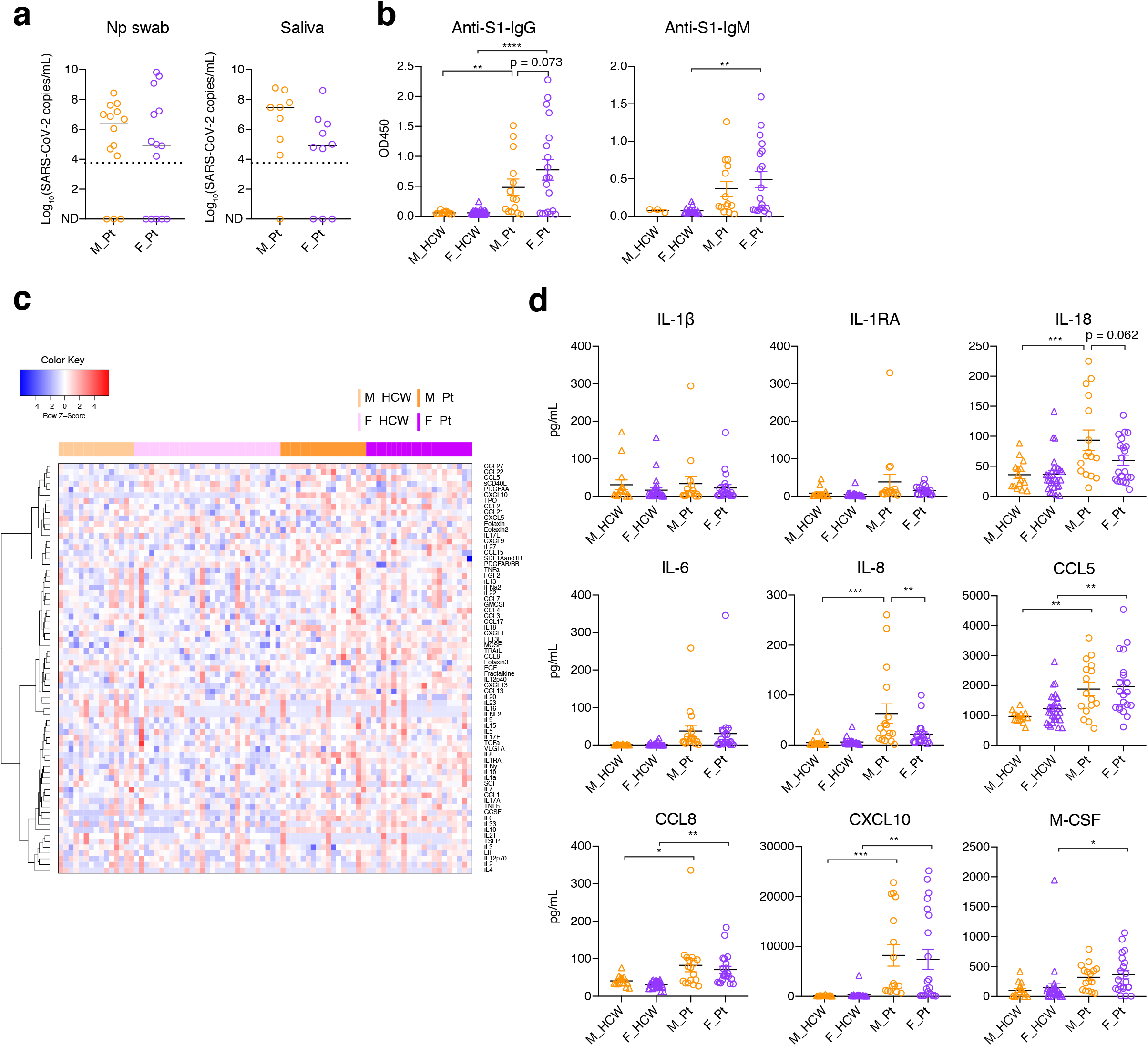
Comparison of viral load, anti-SARS-CoV-2 antibody titers, and plasma cytokines. **a**, Comparison of viral load measured with nasopharyngeal (Np) swab and saliva. Male patients (M_Pt); Female patients (F_Pt) =14:14 for Np samples and 9:10 for saliva samples. Dotted lines indicate the detection limit of the assay (5610 copies/mL), and negatively tested data are shown on the x-axis (not detected; ND). Bar = median. **b**, Titers of specific IgG and IgM antibody against SARS-CoV-2 S1 protein were measured. M_H-CW:F_HCW:M_Pt:F_Pt = 12:74:13:19 for IgG and 3:18:14:19 for IgM. **c**, A heatmap of the plasma levels of 71 cytokines and chemokines in 44 HCW controls (M : F = 15 : 29) and 38 patients in Cohort A (M : F = 17 : 21). Representative innate immune cytokines and chemokines are shown in (**d**). Data = mean ± SEM for **b** and **d**. Bonferroni’s multiple comparison test was used for the comparison between M_Pt vs F_Pt, M_HCW vs F_HCW, M_HCW vs M_Pt, and F_HCW vs F_Pt. *P < 0.05, **P < 0.01, ***P < 0.001, ****P < 0.0001. All p-values < 0.10 are shown. The results of the all the cytokines/chemokines including those shown here can be found in Extended Data Fig. 1 (cytokines/IFNs) and Extended Data Fig. 2 (chemokines/growth factors).

Next, we analyzed the levels of 71 cytokines and chemokines in the plasma. It has been previously reported that levels of many pro-inflammatory cytokines, chemokines and growth factors, including IL-1β, IL-6, IL-8, TNF, CCL2, CXCL10, CCL3, G-CSF and GM-CSF, are elevated in the plasma of COVID-19 patients reflecting innate immune activation^15^, but no studies have thus far analyzed sex differences in cytokine and chemokine levels. In line with previous reports, inflammatory cytokine/chemokine levels were generally higher in patients compared with controls (Fig. 1c). Type-I/II/III interferon (IFN) levels were comparable between sexes in Cohort A (Extended Data Fig. 1). However, we found significantly higher IFNα2 levels in female patients than male patients in Cohort B (Extended Data Table 4 and 5). Levels of many cytokines and chemokines, such as IL-6, CXCL10 (IP-10), CCL5 (RANTES), CCL8, were upregulated in both men and women and the levels between sexes were comparable (Fig. 1d and Extended Data Fig. 1 and 2). However, IL-8 was significantly higher in male patients compared to female patients, and IL-18 was elevated in male patients compared to female patients, albeit not significantly (Fig. 1d). In Cohort B, although we did not see significant sex differences in IL-8 and IL-18, we found significantly higher levels of CCL5 in male patients (Extended Data Table 4). CXCL10 levels were also higher in male patients, albeit not significantly (differences-in-differences, Extended Data Table 5). These data indicated that innate inflammatory cytokines and chemokines are more robustly elevated early and throughout disease course in male patients over female patients.

**Fig. 2.**
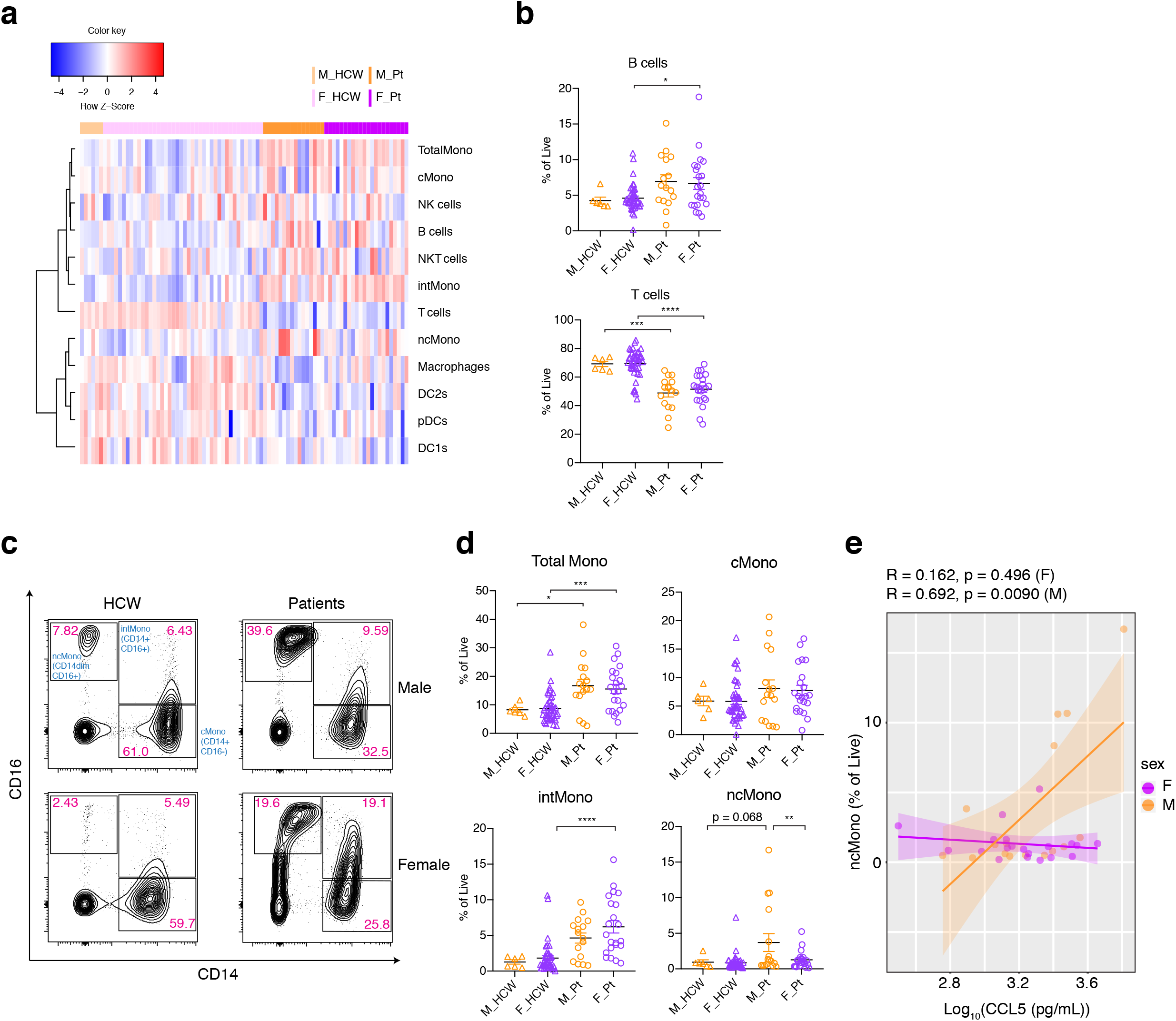
PBMC composition differences between male and female COVID-19 patients. **a**, A heatmap for the composition of PBMC (% of live cells) in control HCWs and Cohort A patients. M_HCW : F_HCW : M_Pt : F_Pt = 6 : 44 : 17 : 22. **b**, Comparison on the proportion of B cells and T cells in live PBMCs are summarized. **c**, Representative 2D plots for CD14 and CD16 in monocytes gate (live/CD19-CD3-/CD56-CD66b-). Numbers in red indicate the percentages in the monocyte gate. **d**, Percentages of total Monocytes, cMono, intMono, ncMono in the total live cells are shown. **e**, Correlation between plasma CCL5 levels and and ncMono (% of live cells). Linear regression line and 95% confidence interval are shown for each sex. Pearson correlation coefficient (R) and p-values for each sex are shown on top of the plot. (**b, d**) Data are mean ± SEM. Bonfer-roni’s multiple comparison test was used for the comparison between M_Pt vs F_Pt, M_HCW vs F_HCW, M_HCW vs M_Pt, and F_HCW vs F_Pt. *P < 0.05, **P < 0.01, ***P < 0.001, ****P < 0.0001. All p-values < 0.10 are shown in the panels.

## Phenotypic differences in monocytes between male and female COVID-19 patients

Next, we examined the immune cell phenotype by flow cytometry. Freshly isolated PBMCs were stained with specific antibodies to identify T cells, B cells, NK-T cells, NK cells, monocytes, macrophages, and dendritic cells (Fig. 2a). As previously reported^16^, T cells were significantly reduced in patients, but the levels were comparable between sexes (Fig. 2a,b), while the proportion of B cells was increased in the patients to levels comparable between sexes (Fig. 2a,b). Notably, we found an increase in total monocyte population in both sexes (Fig. 2a). While CD14^+^CD16^−^ classical monocytes (cMono) were comparable across all groups, levels of CD14^+^CD16^+^ intermediate monocytes (intMono) were elevated in patients compared with HCWs, and this elevation was more robust in female patients (Fig. 2c,d). In contrast, male patients had higher levels of CD14^lo^CD16^+^ non-classical monocytes (ncMono) compared to both controls and female patients (Fig. 2c,d). In cohort B, we also observed higher levels of ncMonos in male patients compared to controls, but no significant difference between sexes (Extended Data Table 5). Since IL-8 (Cohort A) and CCL5 (Cohort B) levels were significantly higher in male patients than in female patients, we examined this elevation was associated with elevation of ncMono in male patients. There was no significant correlation in either sexes of IL-8 (data not shown). However, we found a significant correlation between CCL5 levels and abundance in ncMono only in male patients (Fig. 2e). These findings suggested that progression from classical to non-classical monocytes is arrested at the intermediate stage in female patients, and indicated that elevated innate inflammatory cytokines and chemokines are associated with more robust activation of innate immune cells in male patients.

## Female COVID-19 patients induce more robust T cell response than male patients

We further examined T cell phenotype in COVID-19 patients. The composition of overall CD4-positive cells and CD8-positive cells among T cells were similar between all groups (Fig. 3a, b). Detailed phenotyping of T cells for naïve T cells, central/effector memory T cells (Tcm/Tem), follicular T cells (Tfh), regulatory T cells (Treg) revealed no remarkable differences in the frequency of these subsets between sexes (Fig. 3a). However, we observed more robust induction of CD38 and HLA-DR-positive activated T cells in female patients compared to male patients (Fig. 3a,c,d). In parallel, PD-1 and TIM-3-positive terminally differentiated T cells were more robustly induced in female patients compared to male patients (Fig. 3a,e,f). These findings were seen both in CD4 and CD8 T cells, but the differences were more exaggerated in CD8 T cells (Fig. 3d, f). We also stained for intracellular cytokines in T cells such as IFNγ, Granzyme B (GzB), TNF, IL-6, and IL-2 for CD8 T cells, and IFNγ, TNF, IL-17A, IL-6, and IL-2 for CD4 T cells. Levels of these cytokines were higher in patients compared to controls, and were generally comparable between sexes in the patients (Fig. 3g). Analyses of T cell phenotype in Cohort B did not reveal significant differences between sexes (Extended Data Table 4, 5). Thus, female COVID-19 patients had more abundant activated and terminally differentiated T cell population than male patients at baseline.

**Fig. 3.**
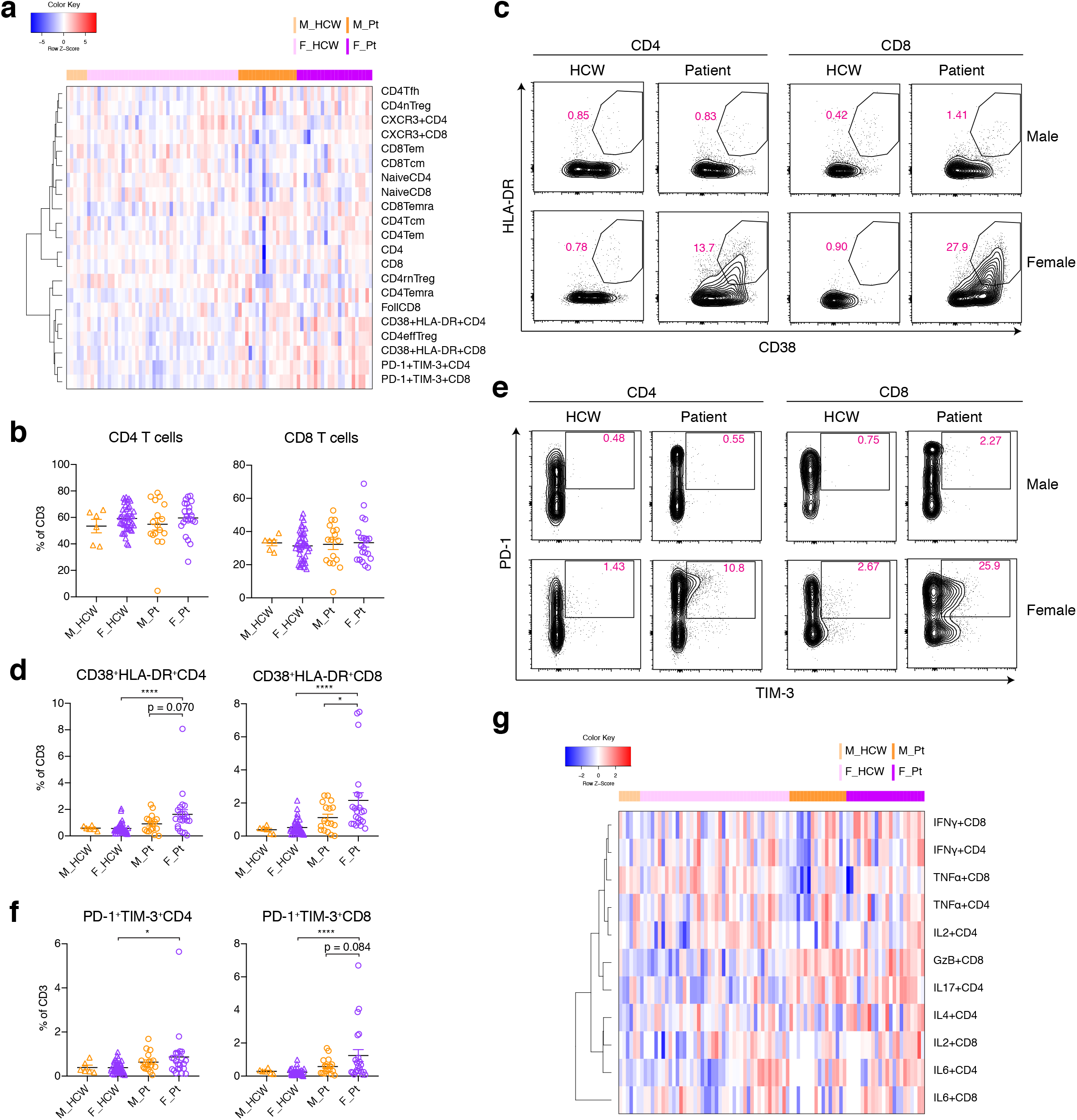
Sex difference in T cell phenotype in COVID-19 patients. **a**, A heatmap for T cell subsets (% of CD3-positive cells) in control HCWs and Cohort A patients. M_HCW : F_HCW : M_PT : F_PT = 6 : 44 : 17 : 22. **b**, Percentages of CD4 and CD8 in the CD3-positive cells are shown. **c**, Representative 2D plots for CD38 and HLA-DR in the CD4 and CD8 T cells are shown. **d**, Percentages of CD38+HLA-DR+ CD4/CD8 cells in CD3-positive cells are summarized. **e**, Representative 2D plots for TIM-3 and PD-1 in the CD4 and CD8 T cells are shown. Data from the same samples in **c. f**, Percentages of PD-1+TIM-3+ CD4/8 cells in CD3-positive cells are summarized. **g**, A heatmap of intracellular cytokine staining of T cells (% of CD3-positive cells). Mean ± SEM in **b, d** and **f**. Bonferroni’s multiple comparison test was used for the comparison between M_Pt vs F_Pt, M_HCW vs F_HCW, M_HCW vs M_Pt, and F_HCW vs F_Pt. *P < 0.05, **P < 0.01, ***P < 0.001, ****P < 0.0001. All p-values < 0.10 are shown in the panels.

## Sex-differences in immune phenotype associated with worsening of COVID-19 disease

Finally, we investigated if certain immune phenotypes and their related factors could be predictive of the severity of the disease, and whether these phenotypes and factors could be different between sexes. To this end, we evaluated the trajectory of the clinical scores of each patient in Cohort A. We then divided the patients in two groups, stabilized group and deteriorated group. The patient was categorized into the deteriorated group if the patient marked a worse, or higher, clinical score at any point compared to the patient’s initial clinical score (see Extended Data Table 3. C_max_ > C_1_), and were otherwise categorized as stabilized.

We first examined the demographics of the 4 groups, namely, stabilized male patients (M_stabilized), deteriorated male patients (M_deteriorated), stabilized female patients (F_stabilized), and deteriorated female patients (F_deteriorated), and found that while M_deteriorated group were significantly older compared with M_stabilized group, the two female groups were comparable in age (Fig. 4a). In addition, body mass index (BMI) for M_deteriorated was higher in the M_stabilized group, while there was no difference between F_deteriorated and F_stabilized groups (Fig. 4a). In contrast, F_stabilized group had lower median values for virus RNA concentrations compared to F_deteriorated group or with two male groups, particularly in saliva (Fig. 4a). We observed higher anti-S IgG levels in the women who had stable disease compared to women with progressive disease, the latter being comparable to men with stable disease or worse disease (Fig. 4a). Thus, the robust anti-S IgG levels in women was associated with their ability to control disease progression.

**Fig. 4.**
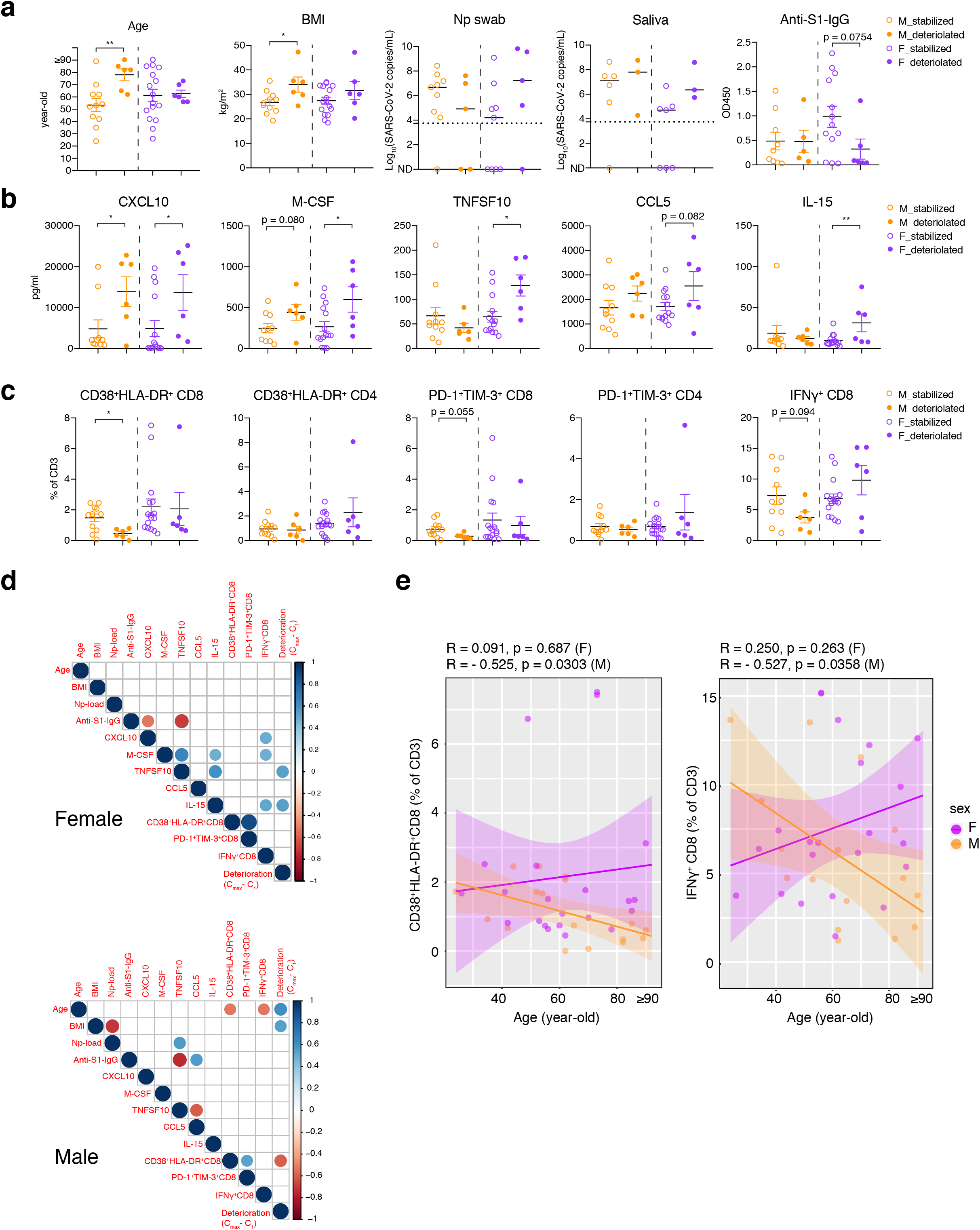
Differential immune phenotypes related to COVID-19 disease progression between sexes. Patients in Cohort A are divided into stabilized group and deteriorated group, depending on the comparison between maximum clinical score after sampling and the score at the sampling (M_stablized ; M_deteriorated : F_stablized : F_aggravated= 11:6:16:6). **a**, The differences in the patients’ age, BMI, nasopharyngeal/saliva virus RNA copies, and anti-S1-IgG antibody are compared. For virus concentration panels, dotted lines indicate the detection limit, and median values are indicated for each group. **b**, Cytokine/chemokine comparison between stabilized and deteriorated group. **c**, Proportions of activated (CD38+HLA-DR+) and terminally differentiated (PD-1+TIM-3+) CD4/CD8 T cells, and IFNγ+CD8 T cells in CD3-positive T cells are shown. **d**, Pearson correlation heatmaps of the indicated parameters are shown for each sex. For viral load levels and cytokine/chemokine levels, log-transformed values were used for the calculation of the correlations. The size and color of the circles indicate the correlation coefficient (R), and only statistically significant correlations (p < 0.05) are shown. Clinical deterioration from the first time point is scored by C_max_-C_1_. **e**, Correlation between age and CD38+HLA-DR+ CD8 T cells (left) and IFNγ+CD8 T cells (right, both in % of CD3 T cells). Linear regression lines and 95% confidence intervals are shown. Pearson correlation coefficient (R) and p-values for each correlation and for each sex are shown on top of each plot. Data are mean ± SEM and unpaired t-test was used to compare the differences between stabilized group and deteriorated group about each sex in **a, b, c**. For the age panel in **a** and correlation plots for age and T cells (**e**), data points for individuals ≥ 90-year-old are plotted as 90-year-old. *P < 0.05, **P < 0.01. All p-values < 0.10 are shown in the panels.

We further investigated if the key factors identified in the previous analyses correlate with disease progression in male and female patients. We observed that regardless of sex, some chemokines and growth factors, such as CXCL10 (IP-10) and M-CSF, were elevated in patients that went on to develop worse disease. However, there were some key innate immune cytokines, such as CCL5, TNFSF10 (TRAIL) and IL-15, which were specifically elevated only in female patients that subsequently progressed to worse disease, but this difference was not observed in male patients (Fig. 4b). In the age-adjusted analysis of Cohort A, we also found that CXCL10 was only elevated in female patients that progressed to worse disease compared to stabilized patients, but no such correlation as found in men (Extended Data Table 6).

T cell phenotypes in these groups revealed that male patients whose disease worsened had a significantly lower proportion of activated T cell population (CD38^+^HLA-DR^+^), and tendencies for fewer terminally differentiated T cell population (PD-1^+^TIM-3^+^) and IFNγ+ CD8 T cells at the first sample collection, compared with their counterpart men who progressed to worse disease (Fig. 4c). However, in women, the deteriorated group had similar levels of these types of CD8 T cells compared with the stabilized group (Fig. 4c).

We finally examined the correlations between age, BMI, viral loads, anti-S1 antibodies, cytokines/chemokines, activated/terminally differentiated/IFNγ-producing CD8 T cells, and clinical disease course (C_max_ – C_1_ was used for the deterioration score). The correlation matrix clearly revealed that in women, higher innate immune cytokines such as TNFSF10 and IL-15 were positively correlated with disease progression, while there was no association between CD8 T cell status and deterioration (Fig. 4d, results of age-adjusted analysis in Extended Data Table 6). In particular, CXCL10 and IL-15 were positively correlated with IFNγ^+^CD8 T cells in female patients (Fig. 4d). Additionally, CXCL10 was negatively correlated with anti-S IgG levels in female patients but not in male patients.

In contrast, in male patients, progressive disease was clearly associated with higher age, higher BMI, and poor CD8 T cell activation (Fig. 4d). Poor CD8 T cell activation and poor IFNγ production by CD8 T cells were significantly correlated with patients’ age, while these correlations were not seen in female patients (Fig. 4d,e). These differences seemed to highlight the differences between the sexes in the immune response against SARS-CoV-2 as well as the difference of the potential prognostic/predictive factors for clinical deterioration of COVID-19.

## Discussion

Our results revealed key differences in immune responses during the early phase of SARS-CoV-2 infection in male and female patients. First, we found that the levels of several important proinflammatory innate immune cytokines such as IL-8 and CCL5 were higher in male patients, which correlated with the robust increase in non-classical monocytes. Second, T cell responses were more robustly activated in female patients compared to male patients. In particular, activated CD8 T cells were significantly elevated only in female patients but not in male patients over healthy volunteers. Analysis of their clinical trajectory revealed that, while poor T cell responses were associated with future progression of disease in male patients, higher innate immune cytokine levels were associated with worsening of COVID-19 disease in female patients. Importantly, the T cell response was significantly and negatively correlated with patients’ age in male, but not in female, patients. These data indicate key differences in the baseline immune capabilities in men and women during the early phase of SARS-COV-2 infection, and suggest a potential immunological underpinning of the distinct mechanisms of disease progression between sexes. These analyses also provide a potential basis for taking sex-dependent approaches to prognosis, prevention, care, and therapy for patient with COVID-19.

Most patients with severe COVID-19 exhibit substantially elevated plasma levels of pro-inflammatory cytokines including IL-6 and IL-1β, as well as IL-2, IL-8, IL-17, G-CSF, GM-CSF, CXCL10, CCL2, CCL3 and TNF, characterized as cytokine storm^15,17^. Activation of these cytokines may lead to shock and tissue damage and subsequently to multiple organ failure.

They also mediate extensive pulmonary pathology, typically presenting with massive infiltration of neutrophils, leading to diffuse alveolar damage^18^. Certain clinical inflammation markers, such as CRP and procalcitonin, which are known to correlate with IL-6 and IL-8 levels, are higher in men in severe cases of COVID-19 (Ref. ^4^). In the present study, we found the higher levels of several cytokines, IL-8, IL-18 (Cohort A), and CCL5 (Cohort B), in the plasma of male patients compared to female patients. A recently published study demonstrated that higher plasma levels of IL-8 are significantly correlated with the decrease in lymphocytes, and lymphopenia (especially the decrease in T cells) is predictive of COVID-19 disease progression^16^. IL-8 is a key chemotactic factor for neutrophils^19,20^, and neutrophils are associated with poorer outcomes in patients with COVID-19 (Ref.^21^). We found robust increase in ncMono in the male patients compared with female patients, while female patients had elevated intMono. ncMono patrol the endothelium in search of injury, and secrete inflammatory cytokines in response to infection, activate endothelial cells, and support the secondary recruitment of neutrophils^22,23^.

In contrast to the higher levels of innate cytokines, we found poor T cell activation in males with less induction of CD8 cells expressing CD38 and HLA-DR, two classical T cell activation markers. Studies have shown that this population exhibits high effector functions, such as proliferation, cytotoxicity, and cytokine production, and this subpopulation of activated CD8^+^ T cells performs an important function during acute viral infections, contributing to viral control^24-27^. In fact, in our cohort, the male patients who progressed in disease severity had lower levels of CD38^+^HLA-DR^+^ CD8 T cells at the first collection. In male patients, loss of activated T cells correlated with old age. In female patients, even older patients were able to develop robust T cell responses. These findings are in line with numerous previous studies that T cells in women are generally more robust compared to men in response to stimulation^9, 28^. The basis of this difference has been at least partially attributed to the fact that many anti-viral genes or inflammatory genes have estrogen response elements (EREs) in their promoter regions^28^.

And of particular importance, we found that activation as well as IFNγ expression of CD8 T cells significantly decline along with age in male patients, while this correlation was not observed in female patients. Age-associated CD8 T cell decrease and dysfunctions have been widely studied and demonstrated^29^, and a recent study has reported the sexual-dimorphism in human immune system aging, including the T cell numbers and phenotypes^30^. Thus, these results collectively paint a picture that in male patients, aging results in T cell activation defects.

Elevated IL-8 levels correlate with lymphopenia^16^. Simultaneously, CCL5 levels were elevated in male patients, potentially leading to the induction and recruitment of inflammatory monocytes to the lung fueling further recruitment of neutrophils, and our finding on the correlation between CCL5 and ncMono abundance is in consistent with this notion. Ultimately, T cell dysfunction with age leads to worse COVID-19 disease outcome. In contrast, in female patients, aging does not appear to lead to T cell dysfunction during COVID-19. Even older women generate robust T cell immunity. However, in some women infected with SARS-CoV-2, they fail to regulate innate immune activation, leading to higher levels of inflammatory cytokines and chemokines, which is associated with worse disease outcome. Female patients that develop anti-S IgG during early infection can suppress proinflammatory cytokines such as CXCL10 and do not progress to worse disease.

Collectively, these data suggest that vaccines and therapies to elevate T cell immune response to SARS-CoV-2 might be warranted for male patients, while female patients might benefit from therapies that dampen innate immune activation early during disease. In summary, our data collectively suggest that the immune landscape in COVID-19 patients is considerably different between the sexes, and these differences may underlie heightened disease susceptibility in men.

## Methods

### Ethics statement

This study was approved by Yale Human Research Protection Program Institutional Review Boards (FWA00002571, Protocol ID. 2000027690). Informed consents was obtained from all enrolled patients and healthcare workers.

### Patients

Adult patients (≥ 18 years old) admitted to Yale-New Haven Hospital between March 18th and May 9th, 2020, positive for SARS-CoV-2 by RT-PCR from nasopharyngeal and/or oropharyngeal swabs, and able to provide informed consent (surrogate consent accepted) were eligible for the Yale IMPACT Biorepository study, and 198 patients were enrolled in this period. Among these patients, we could obtain whole blood for flow cytometry analysis using fresh PBMCs, plasma for cytokine/chemokine measurements, anti-S1 antibody measurements and nasopharyngeal swab and saliva from total of 93 individuals for the present study. For longitudinal analyses, biospecimens (blood, nasopharyngeal swabs, saliva, urine, and/or stool) were collected at study enrollment (baseline) and every 3 to 4 days while in the hospital in 54 of these 93 patients.

The patients were assessed with a locally developed clinical scoring system for disease severity; 1: admitted and observed without supplemental oxygen, 2: required ≤ 3L supplemental oxygen via nasal canal to maintain SpO2 > 92%, 3: received tocilizumab, which per hospital treatment protocol required that the patient to require > 3L supplemental oxygen to maintain SpO2 > 92%, or, required > 2L supplemental oxygen to maintain SpO2 > 92% and had a high sensitivity C-reactive protein (CRP) > 70. 4: the patient required intensive care unit (ICU) level care, 5: the patient required intubation and mechanical ventilation. Detailed demographic information for the entire cohort (93 Cohort B patients, and multiple time point samples from 54 patients among them) and of Cohort A (39 patients) are shown in Extended Data Table 1-3.

### Virus RNA measurement

SARS-CoV-2 RNA concentrations were measured from nasopharyngeal samples and saliva samples by RT-PCR as previously described^31,32^. In short, total nucleic acid was extracted from 300 µl of viral transport media from the nasopharyngeal swab or 300 µl of whole saliva using the MagMAX Viral/Pathogen Nucleic Acid Isolation kit (ThermoFisher Scientific) using a modified protocol and eluted into 75 µl of elution buffer^32^. For SARS-CoV-2 RNA detection, 5 µl of RNA template was tested as previously described^31^, using the US CDC real-time RT-PCR primer/probe sets for 2019-nCoV_N1, 2019-nCoV_N2, and the human RNase P (RP) as an extraction control. Virus RNA copies were quantified using a 10-fold dilution standard curve of RNA transcripts that we previously generated^31^.

### SARS-CoV-2 specific antibody titer measurement

ELISAs were performed as previously described^33^. In short, Triton X-100 and RNase A were added to serum samples at final concentrations of 0.5 % and 0.5 mg/ml respectively and incubated at room temperature (RT) for 30 minutes before use to reduce risk from any potential virus in serum. 96-well MaxiSorp plates (Thermo Scientific #442404) were coated with 50 µl/well of recombinant SARS-CoV-2 S1 protein (ACROBiosystems #S1N-C52H3-100 µg) at a concentration of 2 µg/ml in PBS and were incubated overnight at 4 °C. The coating buffer was removed, and plates were incubated for 1 hour at RT with 200 µl of blocking solution (PBS with 0.1% Tween-20, 3% milk powder). Serum was diluted 1:50 in dilution solution (PBS with 0.1% Tween-20, 1% milk powder) and 100 µl of diluted serum was added for two hours at RT. Plates were washed three times with PBS-T (PBS with 0.1% Tween-20) and 50 µl of HRP anti-Human IgG Antibody (GenScript #A00166, 1:5000) or anti-Human IgM-Peroxidase Antibody (Sigma-Aldrich #A6907, 1:5000) diluted in dilution solution were added to each well. After 1 hour of incubation at RT, plates were washed six times with PBS-T. Plates were developed with 100 µl of TMB Substrate Reagent Set (BD Biosciences #555214) and the reaction was stopped after 12 min by the addition of 100ul of 2 N sulfuric acid. Plates were then read at a wavelength of 450 nm and 570nm.

### Isolation of plasma

Plasma samples were collected after whole blood centrifugation at 400 g for 10 minutes at RT with brake off. The plasma was then carefully transferred to 15 ml conical tubes and then aliquoted and stored at -80 °C for subsequent analysis.

### Cytokine and chemokine measurement

Patients’ sera isolated as above were stored in -80 °C until the measurement of the cytokines. The sera were shipped to Eve technologies (Calgary, Alberta, Canada) on dry ice, and levels of 71 cytokines and chemokines were measured with Human Cytokine Array/Chemokine Array 71-Plex Panel (HD71). All the samples were measured upon the first thaw.

### Isolation of PBMCs

The peripheral blood mononuclear cells (PBMCs) were isolated from heparinized whole blood using Histopaque density gradient under the biosafety level 2+ facility. To isolate PBMCs, blood 1:1 diluted in PBS was layered over in Histopaque in a SepMate tube and centrifuged for 10 minutes at 1200g. The PBMC layer was collected by quickly pouring the content into a new 50ml tube. The cells were washed twice with PBS to remove any remaining histopaque and to remove platelets. The pelleted cells were treated with ACK buffer for red cell lysis and then counted. The percentage viability was estimated using Trypan blue staining.

### Flow cytometry

Exact antibody clones and vendors that were used for flow cytometric analysis are as follows: BB515 anti-HLA-DR (G46-6), BV785 anti-CD16 (3G8), PE-Cy7 anti-CD14 (HCD14), BV605 anti-CD3 (UCHT1), BV711 anti-CD19 (SJ25C1), BV421 anti-CD11c (3.9), AlexaFluor647 anti-CD1c (L161), Biotin anti-CD141 (M80), PE anti-CD304 (12C2), APCFire750 anti-CD11b (ICRF44), PerCP/Cy5.5 anti-CD66b (G10F5), BV785 anti-CD4 (SK3), APCFire750 or PE-Cy7 or BV711 anti-CD8 (SK1), BV421 anti-CCR7 (G043H7), AlexaFluor 700 anti-CD45RA (HI100), PE anti-PD1 (EH12.2H7), APC anti-TIM3 (F38-2E2), BV711 anti-CD38 (HIT2), BB700 anti-CXCR5 (RF8B2), PE-Cy7 anti-CD127 (HIL-7R-M21), PE-CF594 anti-CD25 (BC96), BV711 anti-CD127 (HIL-7R-M21), BV421 anti-IL17a (N49-653), AlexaFluor 700 anti-TNFa (MAb11), PE or APC/Fire750 anti-IFNy (4S.B3), FITC anti-GranzymeB (GB11), AlexaFluor 647 anti-IL4 (8D4-8), BB700 anti-CD183/CXCR3 (1C6/CXCR3), PE-Cy7 anti-IL-6 (MQ2-13A5), PE anti-IL-2 (5344.111), BV785 anti-CD19 (SJ25C1), BV421 anti-CD138 (MI15), AlexaFluor700 anti-CD20 (2H7), AlexaFluor 647 anti-CD27 (M-T271), PE/Dazzle594 anti-IgD (IA6-2), PE-Cy7 anti-CD86 (IT2.2), APC/Fire750 anti-IgM (MHM-88), BV605 anti-CD24 (M1/69), APC/Fire 750 anti-CD10 (HI10a), BV421 anti-CD15 (SSEA-1), AlexaFluor 700 Streptavidin (ThermoFisher). Freshly isolated PBMC were plated at 1-2×10^6^ cells in a 96 well U-bottom plate. Cells were resuspended in Live/Dead Fixable Aqua (ThermoFisher) for 20 minutes at 4°C. Following a wash, cells were then blocked with Human TruStan FcX (BioLegend) for 10 minutes at RT. Cocktails of desired staining antibodies were directly added to this mixture for 30 minutes at RT. For secondary stains, cells were washed and supernatant aspirated; to each cell pellet, a cocktail of secondary markers was added for 30 minutes at 4°C. Prior to analysis, cells were washed and resuspended in 100 μL of 4% PFA for 30 minutes at 4°C. For intracellular cytokine staining following stimulation, cells were resuspended in 200 μL cRPMI (RPMI-1640 supplemented with 10% FBS, 2 mM L-glutamine, 100 U/ml penicillin, and 100 mg/ml streptomycin, X Sodium Pyruvate, and X 2-Mercaptoethanol) and stored at 4°C overnight. Subsequently, these cells were washed and stimulated with 1X Cell Stimulation Cocktail (eBioscience) in 200 μL cRPMI for 1 hour at 37°C. Directly to this, 50 μL of 5X Stimulation Cocktail (plus protein transport inhibitor) (eBioscience) was added for an additional 4 hours of incubation at 37°C. Following stimulation, cells were washed and resuspended in 100 μL of 4% PFA for 30min at 4°C. To quantify intracellular cytokines, these samples were permeabilized with 1X Permeabilization Buffer from the FOXP3/ Transcription Factor Staining Buffer Set (eBioscience) for 10 minutes at 4°C. All further staining cocktails were made in this buffer. Permeabilized cells were then washed and resuspended in a cocktail containing Human TruStan FcX (BioLegend) for 10 minutes at 4°C. Finally, intracellular staining cocktails were directly added to each sample for 1 hour at 4°C. Following this incubation, cells were washed and prepared for analysis on an Attune NXT (ThermoFisher). Data were analysed using FlowJo software version 10.6 software (Tree Star). Set of markers used to identify each subset of cells are summarized in Extended Data Table 7.

### Statistical analysis for the primary analyses

For the primary analyses shown in the main figures, Graph Pad Prism (v8.0) was used for all statistical analysis. Otherwise noted, Bonferroni’s multiple comparison test was used for the comparisons between M_Pt vs F_Pt, M_Pt vs M_HCW, F_Pt vs F_HCW, and M_HCW vs F_HCW for the comparisons. For the comparison between stabilized group and deteriolated group in each sex (Fig. 4 a-c), two-sided unpaired t-test was used for the comparison.

Bioconductor R software was used to generate heatmaps (Fig. 1c, 2a, 3a, 3g), XY graphs for correlation analyses (Fig. 2e and 4e), and Pearson correlation plots (Fig. 4d). For the generation of all heatmaps, log-transformed values were used. For ELISA analysis with Cohort A and HCW samples (n = 82, M_HCW : F_HCW : M_Pt : F_Pt = 15 : 29 : 17 : 21), due to the presence of extreme outliers, the top and bottom 1-percentile values were cut off as the outliers for each cytokine/chemokine, and the remaining values were used for the analyses.

### Statistical analysis for the secondary analyses

All multivariate analyses were conducted using SAS version 9.4 (Cary, NC). We conducted longitudinal analyses of the differences in immune response by sex for patients with COVID-19 and differences in immune response between patients with COVID-19 and healthcare workers by sex and adjusted linear regression to evaluate differences in immune response by sex and patient trajectory.

#### Difference in immune response in COVID-19 positive patients by sex

A marginal linear model was fit to evaluate the difference in various immune responses (outcome) in patients by sex (exposure). We used an auto-regressive correlation structure to account for correlation between repeat observations in an individual over time. To account for the small sample size and unequal follow-up between participants, we used the Morel-Bokossa-Neerchal (MBN) correction. Propensity score methods for covariate adjustment were used to control for the following covariates: age (in years), days since symptom onset (self-reported), ICU status (as a proxy for disease severity) and treatment with either tocilizumab or corticosteroids. A patient was defined as ‘on tocilizumab’ at a given time point if they had received the treatment within fourteen days prior to the time the sample was taken. Patients were defined as ‘on corticosteroids’ if they had received the treatment on the same day the sample was taken. The resulting regression coefficients were interpreted as the difference in immune response between female and male patients.

#### Difference in immune response between COVID-19 positive patients and healthcare workers by sex

To compare differences in immune response between patients and healthcare workers, marginal linear model with a compound symmetric correlation structure and the MBN correction was used to evaluate the difference in immune response between patients and healthcare workers by sex. The model contained terms for sex, study group (patient versus healthcare worker), age (in years) and an interaction term between sex and study group. We calculated the least square means for each group (female patients, female healthcare workers, male patient, male healthcare workers) and evaluated the differences in least square means to compare study groups by sex (female patients vs. female healthcare workers and male patients vs. male healthcare workers). Lastly, the regression coefficient of the interaction term between sex and study group was interpreted as the difference-in-differences between the two groups by sex.

#### Multivariate patient trajectory analysis

We used linear regression to evaluate the difference in baseline immune response between patients who worsened after the baseline sample was taken and those who stabilized by sex. The model contained terms for sex, patient trajectory (worsened vs. stable), age and an interaction term for sex and patient trajectory. We calculated the least square means for each group (female patients who worsened, female patients who stabilized, male patients who worsened and male patients who stabilized) and evaluated the differences in least square means of patient trajectory by sex using the Tukey correction for multiple comparisons. The regression coefficient of the interaction term between sex and patient trajectory was interpreted as the difference-in-differences between the two patient trajectories by sex.

### Yale IMPACT Research Team Authors

(Listed in alphabetical order) Kelly Anastasio, Michael H. Askenase, Maria Batsu, Santos Bermejo, Sean Bickerton, Kristina Brower, Molly L. Bucklin, Staci Cahill, Melissa Campbell, Yiyun Cao, Edward Courchaine, Rupak Datta, Giuseppe DeIuliis, Bertie Geng, Laura Glick, Ryan Handoko, Chaney Kalinich, William Khoury-Hanold, Daniel Kim, Lynda Knaggs, Maxine Kuang, Eriko Kudo, Joseph Lim, Melissa Linehan, Alice Lu-Culligan, Amyn A. Malik, Anjelica Martin, Irene Matos, David McDonald, Maksym Minasyan, M. Catherine Muenker, Nida Naushad, Allison Nelson, Jessica Nouws, Marcella Nunez-Smith, Abeer Obaid, Isabel Ott, Annsea Park, Hong-Jai Park, Xiaohua Peng, Mary Petrone, Sarah Prophet, Harold Rahming, Tyler Rice, Kadi-Ann Rose, Lorenzo Sewanan, Lokesh Sharma, Denise Shepard, Erin Silva, Michael Simonov, Mikhail Smolgovsky, Eric Song, Nicole Sonnert, Yvette Strong, Codruta Todeasa, Jordan Valdez, Sofia Velazquez, Pavithra Vijayakumar, Annie Watkins, Elizabeth B. White, Yexin Yang

## Data Availability

All non PHI data in the manuscript will be made available upon request.

## Acknowledgements

We thank Melissa Linehan for technical and logistical assistance. This work was supported by the Women’s Health Research at Yale Pilot Project Program (AI, AR), Fast Grant from Emergent Ventures at the Mercatus Center, Mathers Foundation, and the Ludwig Family Foundation. A.I. is an Investigator of the Howard Hughes Medical Institute. CBFV is supported by NWO Rubicon 019.181EN.004. AM is supported by NIH grant R37AI041699.

## Author contributions

A.I., S.B.O., A.I.K. conceived the study. C.L., J.K., J.S., J.E.O., T.M. collected and processed patient PBMC samples. B.I., J.K., C.D.O. collected epidemiological and clinical data. F.L., A.M., J.S., E.Y.W., A.R. acquired and analyzed ELISA data. A.L.W., C.B.F.V., I.M.O., R.E., S.L., P.L., A.V., A.P., M.T. performed the virus RNA concentration assays. N.D.G. supervised virus RNA concentration assays. P.W. acquired and analyzed the flow cytometry data. A.C-M., A.J.M. processed and stored patient specimens, J.B.F., C.D.C., and S.F. assisted in patient recruitment, W.L.S. supervised clinical data management. T.T. designed the analysis scheme, analyzed, and interpreted the data for the baseline analyses. M.K.E. and S.B.O. designed the analysis scheme, and interpreted the data for the longitudinal analyses. M.K.E. analyzed the longitudinal data. T.T. and A.I. drafted the manuscript. A.I., A.R., S.B.O. revised the manuscript.

A.I. secured funds and supervised the project.

**Supplementary information** is available for this paper.

## Figure legends

**Fig. 1. Comparison of virus RNA concentrations, anti-SARS-CoV-2 antibody titers, and plasma cytokines at first sampling. a**, Comparison of virus RNA measured from nasopharyngeal (Np) swab and saliva. Male patients (M_Pt); Female patients (F_Pt) =14:14 for nasopharyngeal samples and 9:10 for saliva samples. Dotted lines indicate the detection limit of the assay (5,610 copies/mL), and negatively tested data are shown on the x-axis (not detected; ND). Bar = median. **b**, Titers of specific IgG and IgM antibody titers against SARS-CoV-2 S1 protein were measured. M_HCW:F_HCW:M_Pt:F_Pt = 12:74:13:19 for IgG and 3:18:14:19 for IgM. **c**, A heatmap of the plasma levels of 71 cytokines and chemokines in 44 HCW controls (M : F = 15 : 29) and 38patients in Cohort A (M : F = 17 : 21). Representative innate immune cytokines and chemokines are shown in (**d**). Bonferroni’s multiple comparison test was used for the comparison between M_Pt vs F_Pt, M_HCW vs F_HCW, M_HCW vs M_Pt, and F_HCW vs F_Pt. *P < 0.05, **P < 0.01, ***P < 0.001, ****P < 0.0001. All p-values < 0.10 are shown. The results of all the cytokines/chemokines including those shown here can be found in Extended Data Fig. 1 (cytokines/IFNs) and Extended Data Fig. 2 (chemokines/growth factors).

**Fig. 2. PBMC composition differences between male and female COVID-19 patients at first sampling. a**, A heatmap for the composition of PBMC (% of live cells) in control HCWs and Cohort A patients. M_HCW : F_HCW : M_Pt : F_Pt = 6 : 44 : 17 : 22. **b**, Comparison on the proportion of B cells and T cells in live PBMCs are summarized. **c**, Representative 2D plots for CD14 and CD16 in monocytes gate (live/CD19-CD3-/CD56-CD66b-). Numbers in red indicate the percentages in the monocyte gate. **d**, Percentages of total Monocytes, cMono, intMono, ncMono in the total live cells are shown. **e**, Correlation between plasma CCL5 levels and and ncMono (% of live cells) is shown. Pearson correlation coefficients (R) and p-values for each sex are shown on top of plot. (**b, d**) Bonferroni’s multiple comparison test was used for the comparison between M_Pt vs F_Pt, M_HCW vs F_HCW, M_HCW vs M_Pt, and F_HCW vs F_Pt. *P < 0.05, **P < 0.01, ***P < 0.001, ****P < 0.0001. All p-values < 0.10 are shown in the panels.

**Fig. 3 Sex difference in T cell phenotype in COVID-19 patients at first sampling. a**, A heatmap for T cell subsets (% of CD3-positive cells) in control HCWs and Cohort A patients. M_HCW : F_HCW : M_PT : F_PT = 6 : 44 : 17 : 22. **b**, Percentages of CD4 and CD8 in the CD3-positive cells are shown. **c**, Representative 2D plots for CD38 and HLA-DR in the CD4 and CD8 T cells are shown. **d**, Percentages of CD38^+^HLA-DR^+^ CD4/CD8 cells in CD3-positive cells are summarized. **e**, Representative 2D plots for TIM-3 and PD-1 in the CD4 and CD8 T cells are shown. Data from the same samples shown in **c. f**, Percentages of PD-1^+^TIM-3^+^ CD4/8 cells in CD3-positive cells are summarized. **g**, A heatmap of intracellular cytokine staining of T cells (% of CD3-positive cells). For **b, d** and **f**, Bonferroni’s multiple comparison test was used for the comparison between M_Pt vs F_Pt, M_HCW vs F_HCW, M_HCW vs M_Pt, and F_HCW vs F_Pt. *P < 0.05, **P < 0.01, ***P < 0.001, ****P < 0.0001. All p-values < 0.10 are shown in the panels.

**Fig. 4. Differential immune phenotypes at first sampling and COVID-19 disease progression between sexes**. Patients in Cohort A were divided into stabilized group and deteriorated group, depending on the comparison between maximum clinical score after sampling and the score at the first sampling (M_stablized ; M_deteriorated : F_stablized : F_aggravated= 11:6:16:6). **a**, The differences in the patients’ age, BMI, nasopharyngeal/saliva virus RNA copies, and anti-S1-IgG antibody are compared. For virus concentration panels, dotted lines indicate the detection limit. **b**, Cytokine/chemokine comparison between stabilized and deteriorated group. **c**, Proportions of activated (CD38^+^HLA-DR^+^) and terminally differentiated (PD-1^+^TIM-3^+^) CD4/CD8 T cells, and IFNγ^+^CD8 T cells in CD3-positive T cells are shown. **d**, Pearson correlation heatmaps of the indicated parameters are shown for each sex. For viral load levels and cytokine/chemokine levels, log-transformed values were used for the calculation of the correlations. The size and color of the circles indicate the correlation coefficient (R), and only statistically significant correlations (p < 0.05) are shown. Clinical deterioration from the first time point is scored by C_max_-C_1_. **e**, Correlation between age and CD38^+^HLA-DR^+^ CD8 T cells (left) and IFNγ^+^CD8 T cells (right, both in % of CD3 T cells) are shown. Pearson correlation coefficient (R) and p-values for each correlation and for each sex are shown on top of each plot. Unpaired t-test was used to compare the differences between stabilized group and deteriorated group about each sex in **a, b, c**. For the age panel in **a** and correlation plots for age and T cells (**e**), data points for individuals ≥90-year-old are plotted as 90-year-old. *P < 0.05, **P < 0.01. All p-values < 0.10 are shown in the panels.

